# Enhancing Mass Vaccination Programs with Queueing Theory and Spatial Optimization

**DOI:** 10.1101/2024.06.14.24308958

**Authors:** Sherrie Xie, Maria Rieders, Srisa Changolkar, Bhaswar B. Bhattacharya, Elvis W. Diaz, Michael Z. Levy, Ricardo Castillo-Neyra

## Abstract

**Background:** Mass vaccination is a cornerstone of public health emergency preparedness and response. However, injudicious placement of vaccination sites can lead to the formation of long waiting lines or *queues*, which discourages individuals from waiting to be vaccinated and may thus jeopardize the achievement of public health targets. Queueing theory offers a framework for modeling queue formation at vaccination sites and its effect on vaccine uptake.

**Methods:** We developed an algorithm that integrates queueing theory within a spatial optimization framework to optimize the placement of mass vaccination sites. The algorithm was built and tested using data from a mass canine rabies vaccination campaign in Arequipa, Peru. We compared expected vaccination coverage and losses from queueing (i.e., attrition) for sites optimized with our queue-conscious algorithm to those obtained from a queue-naive version of the same algorithm.

**Results:** Sites placed by the queue-conscious algorithm resulted in 9-19% less attrition and 1-2% higher vaccination coverage compared to sites placed by the queue-naïve algorithm. Compared to the queue-naïve algorithm, the queue-conscious algorithm favored placing more sites in densely populated areas to offset high arrival volumes, thereby reducing losses due to excessive queueing. These results were not sensitive to misspecification of queueing parameters or relaxation of the constant arrival rate assumption.

**Conclusion:** One should consider losses from queueing to optimally place mass vaccination sites, even when empirically derived queueing parameters are not available. Due to the negative impacts of excessive wait times on participant satisfaction, reducing queueing attrition is also expected to yield downstream benefits and improve vaccination coverage in subsequent mass vaccination campaigns.

## INTRODUCTION

The expeditious and equitable distribution of vaccinations and other health services is a cornerstone of public health emergency preparedness. *Queues*, or waiting lines, result from scarce or misallocated resources and volatility in traffic and service patterns; they can hinder the delivery of critical services and thereby jeopardize the achievement of public health targets. Not only can long queues deter people from waiting to receive essential health services, they can erode individuals’ trust in health systems in certain contexts^1,2^ and can thus discourage participation in future programs. Long wait times was a major structural barrier to testing for COVID-19 during the early phase of the pandemic,^3^ and poor planning in some jurisdictions resulted in people waiting hours at some mass COVID-19 vaccination sites.^4–6^ Moreover, excessive queueing during pandemic emergencies also pose health risks, as long wait times may increase exposure to infectious pathogens.^7^

Queueing theory is a branch of applied mathematics that offers a valuable framework for studying the behaviors and effects of waiting lines or queues.^8^ In brief, queueing models aim to capture how a customer population moves through a queueing system via a series of processes dictated by probabilistic rates: arriving at a service site, receiving service, waiting in a queue if the server is busy, or leaving the queue before service is rendered when waiting times exceed a customer’s willingness to wait. Queueing theory is foundational to operations research and has been applied to many facets of healthcare operations, including the triage process in emergency care departments,^9^ staffing needs in operating rooms,^10^ hospital bed management,^11,12^ and outpatient scheduling.^13^ Additionally, it has been applied to COVID-19 vaccine distribution and capacity planning,^7,14–18^ as well as the containment of disease outbreaks, bioterrorist attacks, and other public health emergencies.^19–22^

Mass dog vaccination campaigns (MDVCs) are held annually in Arequipa, Peru to address the re-emergence of canine rabies in the region;^23^ they have important parallels with early pandemic vaccination and testing programs in that success depends, in part, on strategically placing and optimally allocating resources across a discrete number of fixed-location facility sites.^24^ While the World Health Organization (WHO) and Pan American Health Organization (PAHO) recommends a minimum vaccination coverage of 70-80% sustained over multiple years to achieve control and eventual elimination of rabies, the MDVCs in Arequipa, which have relied on convenient or *ad hoc* placement of fixed-location vaccination sites, have continually fallen short of this goal.^25,26^

We have previously developed a data-driven strategy to optimize the placement of fixed-location MDVC sites and found that spatially optimized vaccination sites improves both overall vaccination coverage and spatial evenness of coverage.^26^ However, optimization that addresses spatial accessibility without considering queueing is likely to result in an uneven volume of arrivals across facility sites, which may result in long waiting lines.^26^ Here, we incorporate queueing theory into our existing spatial optimization framework to improve canine rabies vaccine uptake by accounting for both the spatial accessibility of MDVC sites and losses resulting from dog owners who refuse to wait for service in the face of excessive queue lengths (i.e., *queueing attrition*). We compare the performance of our queue-conscious algorithm to the queue-naive algorithm in terms of expected vaccination coverage and queueing attrition and evaluate the sensitivity of our results to misspecification of queueing parameters and the assumption of a constant arrival rate within our queueing model.

## METHODS

### Estimating the relationship between MDVC participation probability and household distance to the nearest vaccination site

Distance to the nearest vaccination site is an important predictor of a dog owner’s participation in an MDVC,^23,24,26^ and we quantified this relationship using previously described methods.^26^ Briefly, data on household participation in the most recent MDVC were obtained from post-MDVC surveys that were conducted annually in Arequipa, Peru from 2016-2019. Shortest walking distances between surveyed household residences and their nearest MDVC site were obtained using the Mapbox Directions API and Leaflet Routing Machine.^26–28^ We focused on walking distance instead of other distance metrics, because car-ownership rates are low, and transit options are limited in Arequipa. We fit a Poisson regression model that treated MDVC participation as the outcome variable and walking distance to the nearest MDVC site as the predictor variable; we fit the model by constructing 30-meter distance bins and predicting the number of participating households offset by the number of households per bin. We chose Poisson regression over other statistical models, as we have previously found that the Poisson model provided the best fit for these data.^26^

### A queueing model for MDVCs

We modeled queueing, vaccination, and attrition at each MDVC vaccination site according to an M/M/1 system with first-in-first-out (FIFO) service (figure 1). The M/M/1 system is a widely used queueing model for single server systems and assumes that customer arrivals occur according to a Poisson process, and job service times are independent and identically distributed (iid) exponential random variables that are independent of the arrival process and queue length. Applied to MDVCs, the M/M/1 queueing model assumes that dogs arrive with their owners to a vaccination site according to a Poisson process with arrival rate *λ*, meaning that the interarrival times are iid and follow an exponential distribution with parameter *λ*. The service times (*i.e.*, the time it takes for a dog to get vaccinated) are iid exponential with parameter *μ*, such that the average service time is equal to 1/*μ*. The system is assumed to be FIFO, meaning that dogs are vaccinated in the order that their owners join the queue. Only one dog can get vaccinated at a time, as there is only one vaccinator per site, and dogs are assumed to leave the system as soon as they get vaccinated.

**Figure 1.**
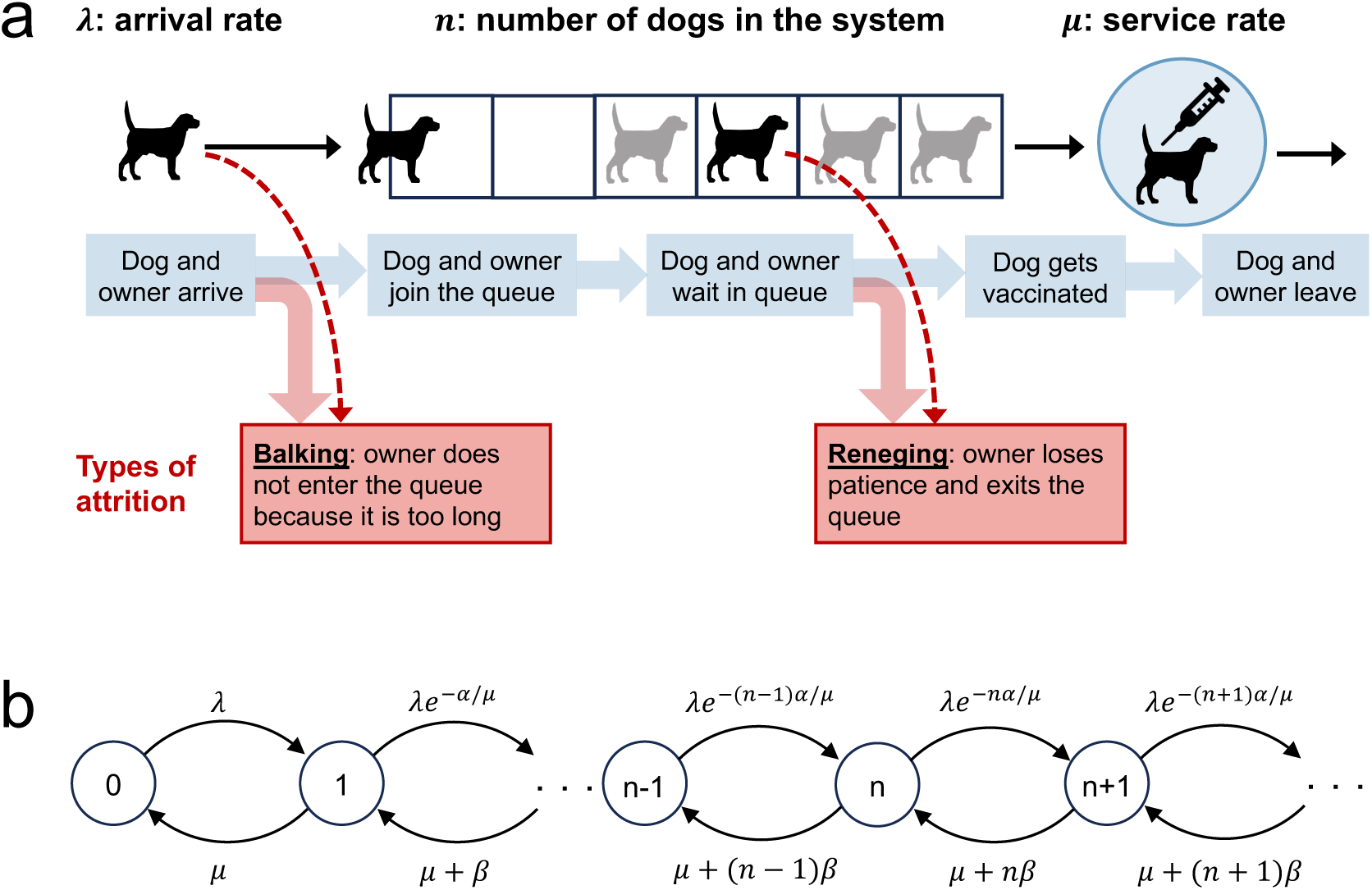
An M/M/1 first-in-first-out queueing model for an MDVC vaccination site. Panel a illustrates the processes captured by the queueing model, with the forms of queuing attrition highlighted by the red boxes. Panel b shows the transition-state diagram for the queueing model, where states, depicted by circles, are defined by the number of dogs in the system, and transitions between states, depicted with curved arrows, are labeled by their corresponding transition rates.

The service rate *μ* was assumed to equal 30 hr^-1^ in accordance with the empirical observation that it takes two minutes on average to vaccinate a dog. The arrival rates were assumed to vary across MDVC sites and were determined as follows. First, the MDVC participation probability function described above was applied to all households falling within an MDVC site’s *catchment* (i.e., all houses closest to the given MDVC site in terms of travel distance) to determine the probability that each household would participate in the MDVC if the house were inhabited and owned dogs. These probabilities were summed and then scaled by the habitability rate, household-dog-ownership rate, and average number of dogs per dog-owning household that were estimated for the study area from post-MDVC surveys (57%, 40%, and 1.86, respectively) to obtain the total number of dogs arriving at MDVC site *s*. This number was then divided by the total operation time for the MDVC site to obtain *λ*_s_, the arrival rate for site *s*.

A dog enters the queueing system at site *s* after it arrives at the site and its owner elects to join the vaccination queue. However, some owners may decline to join the queue if they judge the queue to be too long. This first form of attrition is known as *balking* and was modeled by modifying the arrival rate *λ_s_* so that it decreases by a discouragement factor *e*^-*αn*/*μ*^ < 1.^8^ The *modified arrival rate λ_s,n_* captures the rate that owners join the queue after accounting for those that balk and is given by:

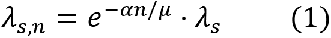

where *n* is the number of dogs that are currently in the system (waiting in queue or being vaccinated), *μ* is equal to the service rate, and *α* is a parameter that scales with balking propensity.^8^

The other form of attrition, known as *reneging*, occurs when an owner who has already joined the queue loses patience and exits the queue before their dogs are vaccinated. We modeled reneging by modifying the service rate *μ* to capture all those leaving the system - both those leaving after vaccination and those who renege. This *exit rate μ_n_* is equal to:

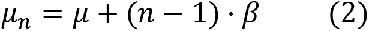

where the second term captures the rate that each of the present *n* – 1 dogs in queue are reneging, and *β* scales with reneging propensity.^8^ Note that in the equations above, the rates of attrition (both balking and reneging) increases with the queue length *n* – 1, as expected.

In order to calculate the expected number of dogs vaccinated during an MDVC, we need to find a closed-form expression for the vaccination rate at a given vaccination site that accounts for losses due to attrition. The derivation of these closed-form equations can be found in the electronic supplementary materials, text A, and are based on the stationary distribution of the queueing model, *i.e.*, on *p*_*s*,*n*_, the probability of finding *n* dogs in the queueing system at MDVC site *s* with arrival rate *λ*_s_:

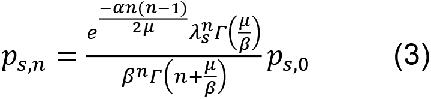

where *Γ*(*z*) denotes the gamma function, i.e., *Γ*(*n*) = (*n* − 1)! for any integer *n* > 0 and *Γ*(*z*) = ∫ *t*^*z*−1^*e*^−*t*^*dt* interpolates the factorial function to non-integer values, and *p*_s,0_ is a normalizing constant given by:

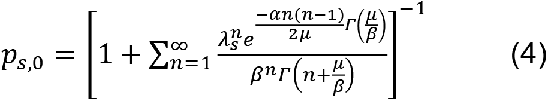

The expected rate that dogs are vaccinated at site *s* is then equal to:

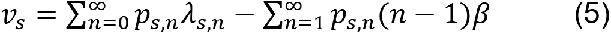

where the first term is equal to the rate that dog owners join the queue after accounting for balking, and the second term is equal to the rate that dog owners renege and thus leave the queue before their dogs are vaccinated. The expected number of dogs vaccinated during an MDVC is thus equal to:

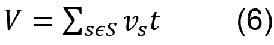

where *S* is the set of all selected vaccination sites and *t* is equal to the total operation time, which is assumed to be the same for all MDVC sites.

In addition to the closed-form equations for the expected behavior of the MDVC queueing system, which were derived assuming the system had reached steady state, we also conducted stochastic simulations to study the behavior of the system in the absence of such assumptions. Simulations were conducted for low- and high-attrition parameter regimes (low: *α* = 0.01 and *β* = 0.02; high: *α* = 0.1 and *β* = 0.1) and for a range of arrival rates (0.5-37.5 dogs/hour in increments of 0.5 dogs/hour). An MDVC site operates for four weekend days (over two weekends) for about four hours per day (*t* = 16 total hours). To mimic these conditions, a single simulation consisted of four independent four-hour-long trials (days), each initialized with no dogs in the queue at time zero; the number of dogs vaccinated each day was summed across the four days to obtain the total dogs vaccinated at an MDVC site. The simulation was run for 1,000 iterations per set of parameter values, and the simulation results were compared to the expected number of dogs vaccinated as determined via the closed-form equations to see how well the two approximated each other.

### Optimizing the locations of vaccination sites

We optimized the placement of MDVC sites for the Alto Selva Alegre district of Arequipa; no more than 20 sites can operate in this region during a campaign due to resource constraints, and 70 locations have been approved by the Ministry of Health for use as feasible MDVC sites (figure 2).^26^ We determined the optimal placement of *k* = 20 sites among these 70 candidate sites by implementing a hybrid recursive interchange-genetic algorithm (electronic supplementary materials, text B and figures S1-S2). The recursive interchange portion of our algorithm is similar to Teitz and Bart’s solution to the *p*-median problem that solves the facility location problem by minimizing the average distance traveled by all households to their nearest site,^29^ but instead of minimizing average walking distance, our algorithm aims to maximize total MDVC participation probability. Maximizing MDVC participation probability allows our algorithm to account for both distance between households and MDVC sites and queue-length-dependent attrition rates.

**Figure 2.**
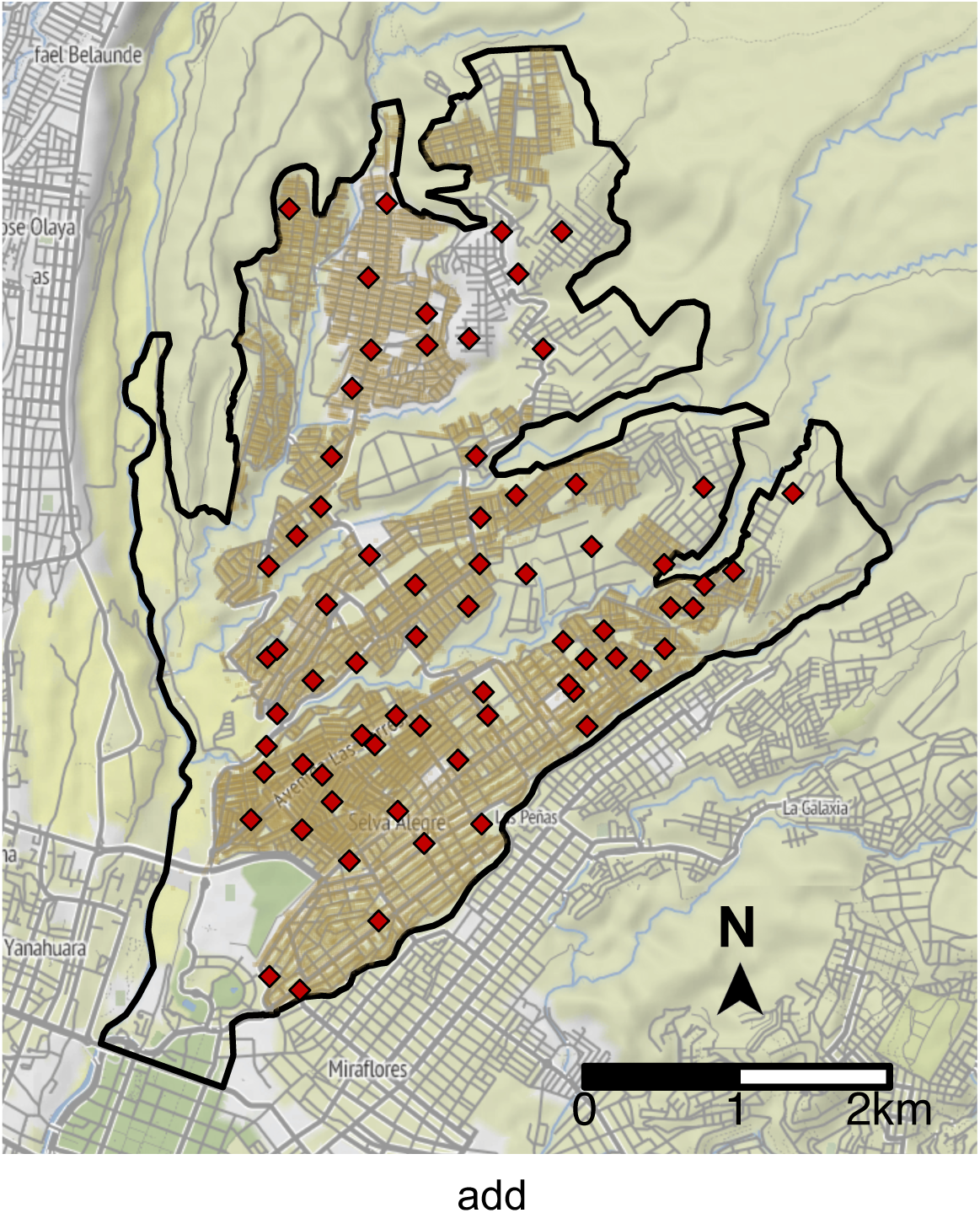
Potential vaccination site locations in Alto Selva Alegre. The boundaries of Alto Selva Alegre are depicted by the solid, black line. Candidate MDVC sites (N = 70) are indicated by red diamonds, and the locations of houses are shaded brown.

The general steps of the recursive interchange algorithm are as follows:

1. Select a random subset of 20 vaccination sites and use the MDVC participation probability function to determine the expected arrival rate *λ* at each site.
2. Calculate the expected number of dogs vaccinated at each site and sum across all sites to calculate the total number of dogs vaccinated.
3. Exchange one selected site with all non-selected candidate locations and keep the one that maximizes the number of dogs vaccinated.
4. Repeat step 3 with remaining sites to obtain a locally optimized set of sites.
5. Perform steps 1-4 over 1,000 iterations, initializing each iteration with a different random subset of sites.

An animation showing a single iteration of the recursive interchange algorithm can be viewed as a video in the electronic supplementary materials. The recursive interchange algorithm was repeated over 1,000 iterations to increase performance, as the algorithm does not guarantee a globally optimal solution. Performance was further enhanced by combining the recursive interchange algorithm with a genetic algorithm that “mates” parental sets output by the recursive interchange algorithm, mimicking natural selection by introducing crossover and mutation and ultimately producing new starting sets on which to repeat the recursive interchange algorithm. The cycling between the recursive interchange and genetic algorithms were repeated until the expected number of dogs vaccinated did not increase over two subsequent rounds of optimization (stopping condition). A full description of the hybrid algorithm can be found in the electronic supplementary materials, text B.

MDVC sites were optimized under three scenarios: no attrition (*α* = *β* = 0), low attrition (*α* = 0.01, *β* = 0.02), and high attrition (*α* = 0.1, *β* = 0.1). Note the no-attrition scenario is the least realistic, as some degree of balking and reneging is expected to occur in the real world. The low- and high-attrition *queue-conscious* solutions were compared to the *queue-naive* solution obtained under the assumption of no attrition (*i.e.,* all dogs that arrive get vaccinated) to determine how the incorporation of queueing behaviors impacted the expected vaccination coverage and the amount of dogs lost to attrition. Note that although the *queue-naive* solution to the location problem was obtained assuming no attrition, its performance was assessed under the assumption of a low- or high-attrition parameter regime. Additionally, the optimized sites were mapped along with their catchments to compare how site placement varied between the queue-conscious and queue-naive solutions.

### Sensitivity analyses

To determine how our results may have been impacted by misspecification of *α* and *β*, we considered four possible scenarios for true balking and reneging propensities. In addition to the low- and high-attrition scenarios discussed previously (*α* = 0.01/*β* = 0.02 and *α* = 0.1/*β* = 0.1, respectively), we considered two additional scenarios for true balking and reneging propensities: (1) low balking and high reneging (*α* = 0.01, *β* = 0.1) and (2) high balking and low reneging (*α* = 0.1, *β* = 0.02). We applied the low- and high-attrition solutions to these four scenarios to evaluate performance (in terms of number of vaccinations and losses to attrition) for situations in which *α* and *β* are correctly and incorrectly specified. For each scenario and queue-conscious solution applied, performance was evaluated using the number vaccinated and losses to attrition achieved by the queue-naive solution as a benchmark.

The optimization methods detailed above rely on the use of the closed-form equations for the queueing system, which assumes a constant arrival rate *λ*. We considered how this assumption impacted our results by allowing *λ* to vary in a step-wise manner to approximate time-varying arrival rates that have been observed in the field (electronic supplementary materials, figure S3). Four time-varying arrival densities were considered: a) a steep unimodal peak density, b) a wide unimodal density that is skewed right, c) a wide unimodal density that is skewed left, and d) a bimodal density distribution (supplementary materials, figure S4). Eight total scenarios were considered, representing all combinations of the four time-varying arrival densities and low- and high-attrition parameter regimes. Queueing simulations were performed for each scenario, and natural splines were used to summarize the behavior of the system over a range of arrival rates (electronic supplementary materials, text C). Once again, the performance of the low- and high-attrition solutions were assessed for each scenario, using performance under the queue-naive solution as a benchmark. Additionally, the different nonconstant arrival rate densities were compared to the baseline assumption of a constant arrival rate to determine how this assumption impacted estimations of the number of vaccinations and losses to attrition.

## RESULTS

### Relationship between MDVC participation probability and household distance to the nearest vaccination site

The probability that a dog-owning household participated in an MDVC was inversely associated with the distance a householder had to walk to the nearest MDVC site (electronic supplementary materials, figure S5). The Poisson regression model determined that a household located in very close proximity to an MDVC site (walking distance < 30 m) had a participation probability of 75% compared to 38% for a household that would have to walk one kilometer to the nearest MDVC site.

### Queue-conscious optimization for MDVCs

As expected, the amount of balking and reneging was greater for higher arrival rates and for higher *α* and *β* values, representing greater attrition propensity (figure 3). Although the closed-form expression for the expected number of vaccinations (equation 6) was derived under steady-state assumptions, the results of the stochastic simulations closely approximated results obtained using equation 6 across a range of arrival rates (electronic supplementary materials, figure S6). Thus, equation 6 was used as the objective function in the hybrid algorithm that was used to optimize MDVC site placement.

**Figure 3.**
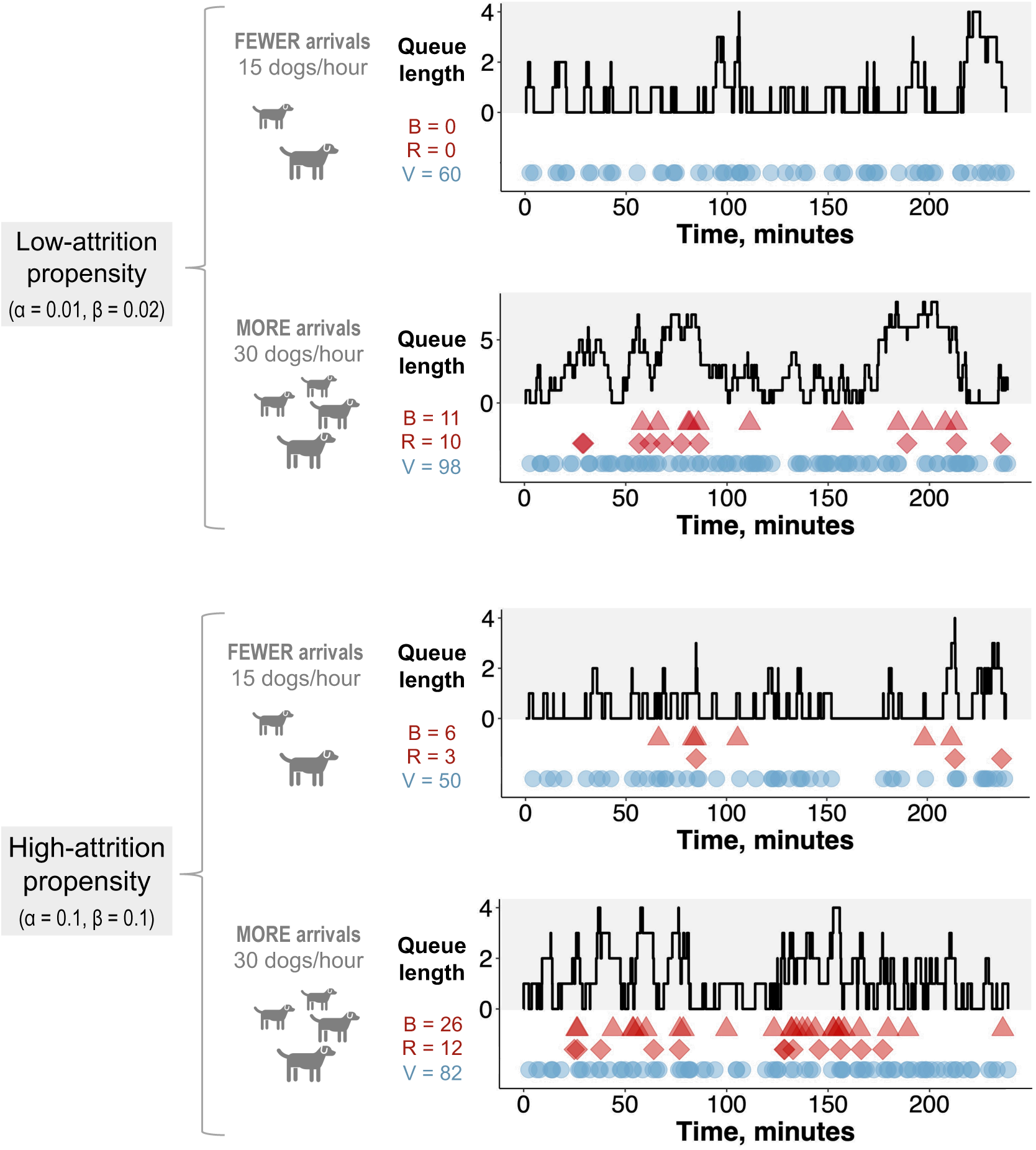
Realized trials of the stochastic queueing model. Each trial of the stochastic queueing simulation represents a single four-hour day at an MDVC site. The gray-shaded portion of each plot tracks the queue length over the four-hour period, and the colored shapes in the white portion of each plot tracks the occurrences of balking (red triangles), reneging (red diamonds) and vaccination (blue circles). The number of balking events (B), reneging events (R), and vaccinations (V) are reported for each trial. Trials are shown for two different α/β parameter regimes (low: *α* = 0.01, *β* = 0.02 and high: *α* = 0.1, *β* = 0.1) and two different arrival rates (15 and 30 dogs per hour).

Compared to the queue-naive algorithm, the queue-conscious algorithm favored a more even distribution in the number of arrivals across all selected sites (figure 4 and electronic supplementary materials, figure S7). The queue-conscious algorithm “flattens” the distribution of arrivals by placing more sites in densely populated areas to divide the higher vaccination workload across more vaccinators and placing fewer sites in less populous areas (figure 5 and electronic supplementary materials, figure S8). This difference in site distribution is expected, because too many arrivals at a site results in the formation of long queues and more losses from balking and reneging; these losses are accounted for (penalized) by the queue-conscious algorithm but not by the queue-naive algorithm, which assumes that all arrivals get vaccinated.

**Figure 4.**
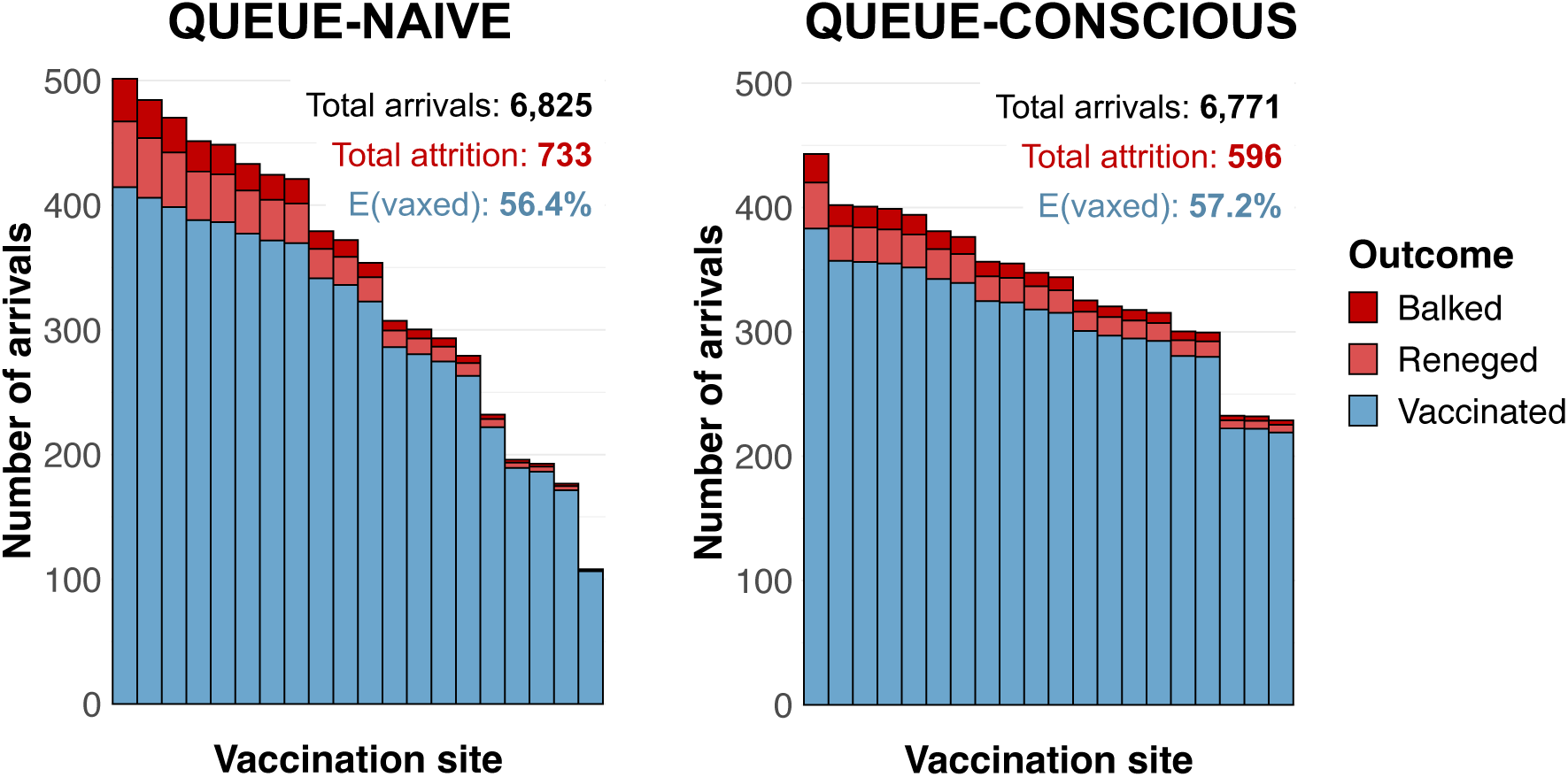
Arrivals histograms for sites selected by queue-naive vs. queue-conscious optimization for the low-attrition system (*α* = 0.01, *β* = 0.02). The height of each stacked bar represents the expected number of dogs that arrive at a selected vaccination site. Bars are subdivided by color according to whether dogs ultimately get vaccinated (blue) or are lost to attrition, either through balking (dark red) or reneging (light red). The text above the bars give the total number of arrivals, total losses to attrition, and overall vaccination coverage achieved for each algorithm. While the queue-naive sites were obtained by the hybrid algorithm without considering attrition, the number of dogs vaccinated and the number of dogs lost to attrition for both queue-naive and queue-conscious sites were determined assuming low-attrition parameter values using equation 6 and the equations outlined in the electronic supplementary materials, text A.

**Figure 5.**
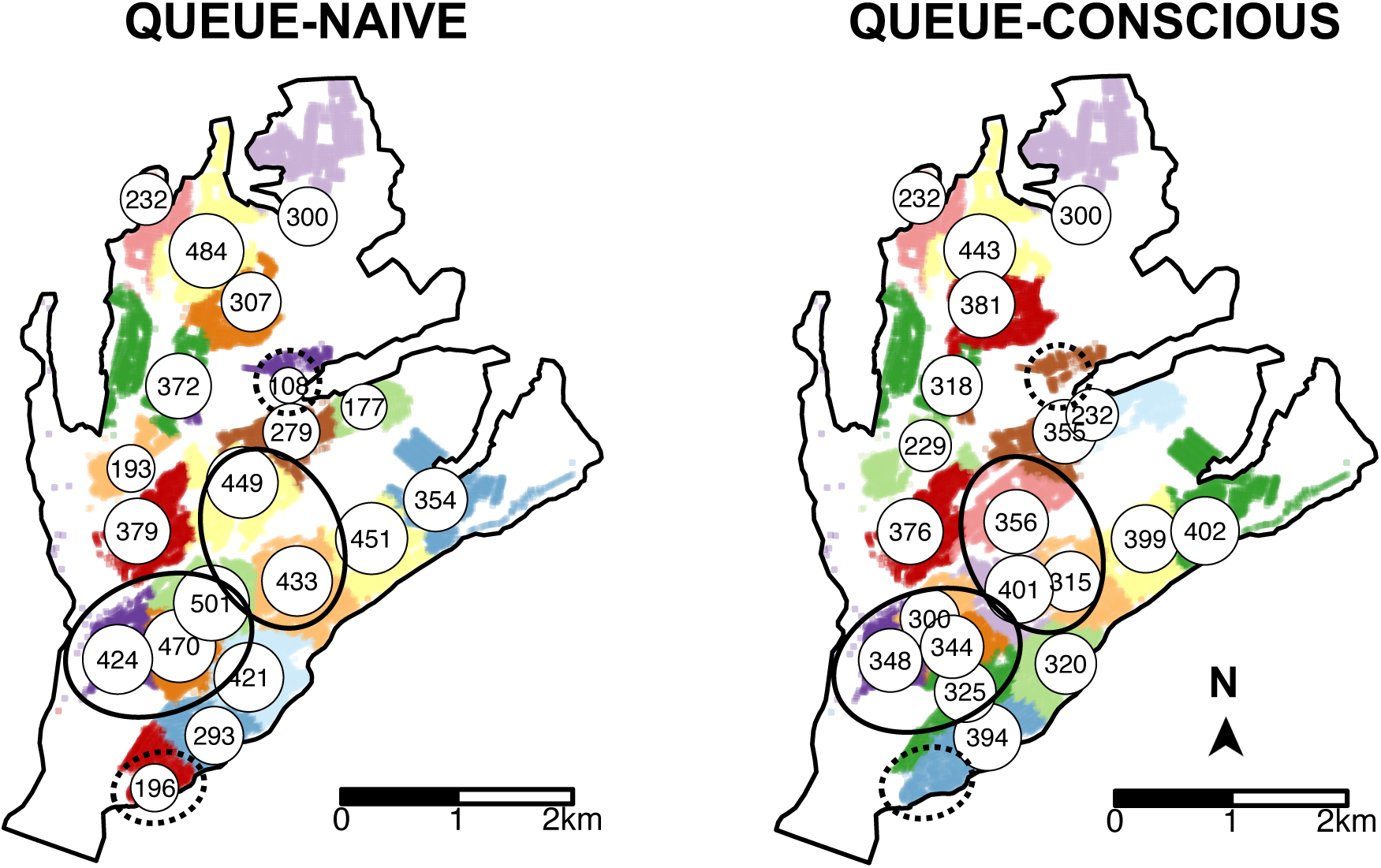
Locations of MDVC sites selected by the queue-naive vs. queue-conscious algorithm for the low-attrition system (*α* = 0.01, *β* = 0.02). The locations of selected vaccination sites are indicated by white circles that are labeled and scaled according to the expected number of arriving dogs, which were calculated using equation 6. Houses in the study area are small dots colored according to their catchment, representing the area in which a MDVC site is the closest site for houses in terms of travel distance. Areas in which the queue-conscious algorithm placed a higher density of vaccination sites compared to the queue-naive algorithm are indicated by ellipses with solid lines, and areas in which the queue-conscious algorithm placed one fewer site are indicated by ellipses with dotted lines.

Within the low-attrition system (*α* = 0.01, *β* = 0.02), vaccination sites that were placed using the queue-conscious algorithm achieved an expected vaccination coverage of 57.2% compared to 56.4% achieved by the queue-naive algorithm (figure 4). The amount of queueing attrition (i.e. the expected number of dogs whose owners balked or reneged) was also lower for sites placed using the queue-conscious algorithm: 596 vs. 733 for the queue-naive algorithm, representing a 19% reduction (figure 4). Trends were similar for the high-attrition system (*α* = 0.1, *β* = 0.1), in which the queue-conscious algorithm improved the expected vaccination coverage from 47.2% to 48% and reduced queueing attrition by 9% from 1,727 to 1,566 (electronic supplementary materials, figure S7).

### Sensitivity analyses

These results were robust to misspecification of *α* and *β*, and the performance varied only slightly between the high- and low-attrition solutions for all combinations of *α* and *β* considered (figure 6). When the true values of *α* and *β* are low (*α* = 0.01 and *β* = 0.02), overestimating these parameters in the optimization did not result in a substantial loss in the number of dogs vaccinated compared to the correctly optimized solution (82 vs. 84 more dogs vaccinated beyond the queue-naive solution). Similarly, when the true values of α and β are high (*α* = β = 0.1), underestimating these parameters in the optimization did not markedly impact the number of dogs vaccinated compared to the correctly optimized solution (83 vs. 85 more dogs vaccinated beyond the queue-naive solution). Moreover, applying the low- and high-attrition solutions resulted in a similar number of dogs vaccinated when the true value of *α* is low and the true value of *β* is high and vice-versa (figure 6a). The high-attrition solution resulted in a greater reduction in queueing attrition than the low-attrition solution for all four attrition scenarios, though both solutions resulted in substantially fewer losses compared to the queue-naive solution (figure 6b)

**Figure 6.**
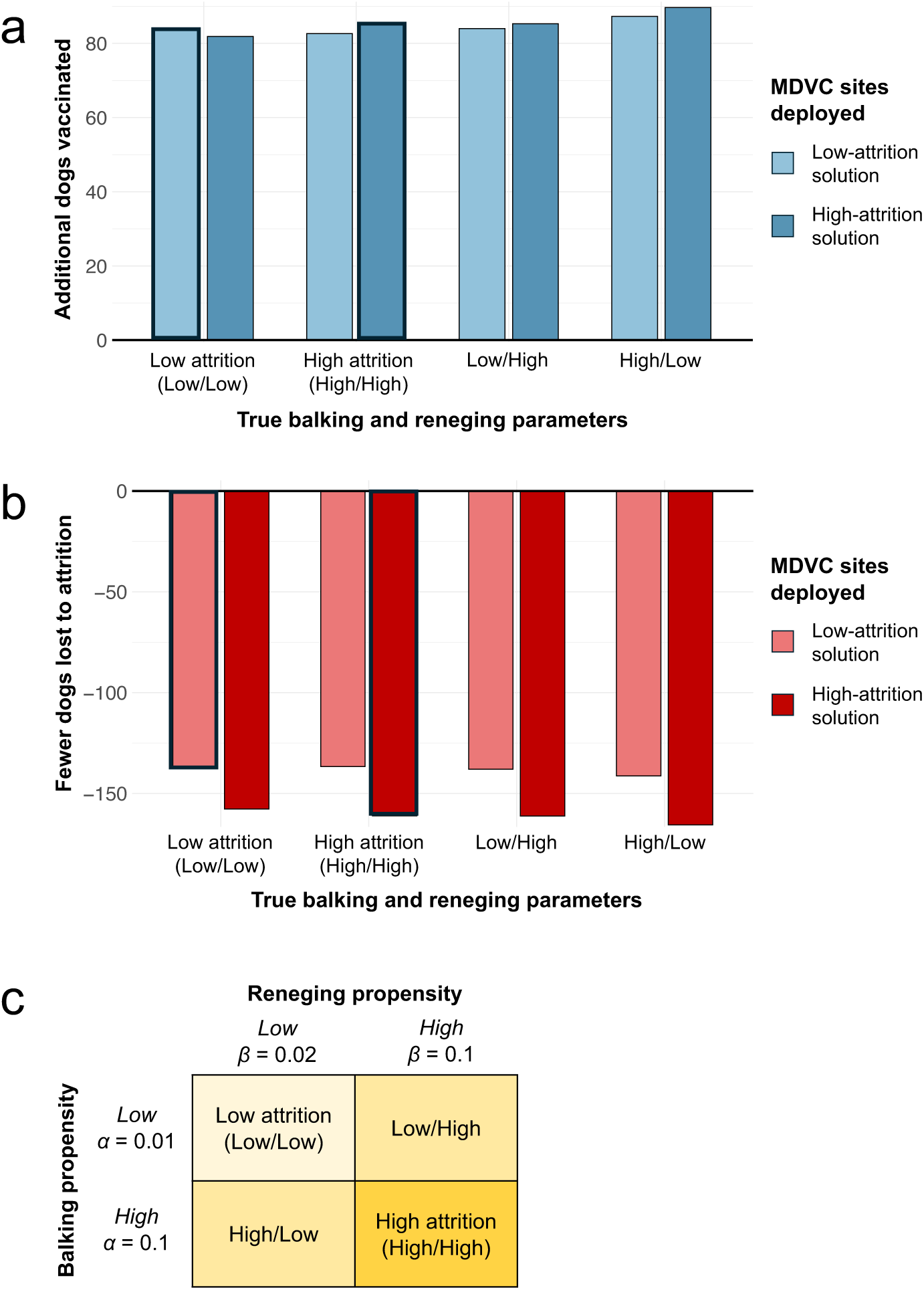
Sensitivity of results to misspecification of balking and reneging parameters. Panels a-b illustrate how misspecification of *α* and *β* impacts the expected number of dogs vaccinated (a) and the number of dogs lost to attrition (b). The performance of the low- and high-attrition solutions are provided with the queue-naive solution acting as a benchmark; thus, (a) shows the additional number of dogs vaccinated beyond the expected number achieved with the queue-naive solution, and (b) shows the reduction in attrition compared to the queue-naive solution. Bars outlined in bold represent scenarios in which the balking and reneging parameters are correctly estimated in the optimization. Panel c provides a legend with the values of *α* and *β* for the four balking/reneging scenarios considered.

The superior performance of the queue-conscious algorithm compared to the queue-naive algorithm was also robust to relaxation of the constant arrival rate assumption. For all four time-varying arrival densities and attrition regimes, both low- and high-attrition solutions substantially outperformed the queue-naive solution in terms of the numbers vaccinated and lost to attrition (electronic supplementary materials, figure S9). Interestingly, with the exception of arrival density D under a low-attrition regime, for which the low- and high-attrition solutions yielded roughly equal numbers of vaccinations, the high-attrition solution outperformed the low-attrition solution in terms of the numbers vaccinated. The high-attrition solution also resulted in less queueing attrition than the low-attrition solution for all scenarios considered. In addition, nonconstant arrival rates resulted in more queueing attrition and fewer dogs vaccinated compared to an otherwise equivalent scenario where the constant arrival rate assumption is met (electronic supplementary materials, figure S10).

## DISCUSSION

We developed an optimization algorithm that integrates queueing theory into a spatial optimization framework to improve the placement of mass vaccination sites. We applied our algorithm to the MDVC in Arequipa, Peru by simultaneously minimizing travel distance to MDVC sites and queueing attrition resulting from large arrival volumes at some sites. Our queue-conscious algorithm decreased queueing attrition by 9-19% and increased expected vaccination coverage by 1-2% compared to a queue-naive version of the same algorithm. MDVC site optimization that accounted for queueing placed more vaccination sites in densely populated areas to even out the number of expected arrivals across sites, and sensitivity analyses revealed that accounting for queueing resulted in improved MDVC performance, even in the absence of accurate parameter estimates. Moreover, the expected gains in vaccination coverage do not capture the indirect effects of excessive wait times and attrition.

Longer wait times have been negatively associated with patient satisfaction in a variety of healthcare contexts, and patients report being less likely to repeatedly patronize a medical practice with long wait times compared to one with shorter wait times.^1,30,31^ In the context of canine rabies vaccination programs, individuals that have to wait a long time before receiving vaccinations for their dogs may be far less likely to participate in subsequent rabies vaccination campaigns. Furthermore, in light of the growing body of literature supporting the social contagion of vaccine hesitancy, vaccine uptake, and participation in public health campaigns,^32–36^ there is also potential for a negative cascade if dog owners who experience long wait times at an MDVC site tell friends and neighbors about their negative experiences and discourage turnout within their social networks. Taken together, the reduction of attrition resulting from well-placed vaccination sites may pay dividends in improving turnout and vaccination coverage in subsequent MDVCs; this is particularly important for canine rabies elimination, which requires sustained high levels of vaccination year after year.^37–39^

We assumed that owners arrived with their dogs to MDVC sites at time-invariant rates. The rationale behind this assumption was two-fold: (1) it ensured tractability of the queueing equations, and (2) it was unclear how to specify a nonconstant arrival rate in the face of heterogeneity in the trajectory of rates observed at MDVC sites (electronic supplementary materials, figure S3). While this simplifying assumption could raise concern about the validity of our results, our sensitivity analysis that probed this assumption indicated that the queue-conscious solutions outperformed the queue-naive solutions even when arrival rates varied over time (electronic supplementary materials, figure S9). We also found that nonconstant arrival rates resulted in more queueing attrition and fewer dogs vaccinated than the baseline assumption of a constant arrival rate (electronic supplementary materials, figure S10). This result is unsurprising because a time-varying arrival density would lead to swells of arrivals during peak intervals, when queue lengths would escalate and cause attrition to spike.

Surprisingly, the high-attrition solution performed as well as or better than the low-attrition solution for all time-varying arrival scenarios, even those in which the true attrition rates were low (electronic supplementary materials, figure S9). This result can be explained by the spikes in attrition that accompany time-varying arrival rates but are not captured by the low-attrition solution, which are obtained under the assumption of a constant arrival rate. As a result, even when *α* and *β* are low, the expected vaccination rate is higher with the high-attrition solution, as it favors a more even distribution in the number of arrivals across vaccination sites (compare figures 4-5 to electronic supplementary materials, figures S7-S8). These results suggest that applying MDVC optimization in the real world is as much an art as it is a precise science. Even if the “true” balking and reneging rates could be determined, it may be beneficial to slightly overestimate these parameters to offset the reality of nonconstant arrival rates.

The queue-conscious algorithm we employed decreases queue lengths across the study area, but some queueing is inevitable. Attrition can be minimized further by improving the waiting experience for queueing dog owners.^40,41^ In the context of MDVCs, accommodations should be made for aggressive dogs, whose presence in a queue can cause other owners to balk or renege. Some vaccinators may choose to deviate from FIFO principles and vaccinate aggressive dogs first regardless of when they arrive to remove them from the queue more quickly. This priority service approach (where aggressive dogs take priority over less aggressive dogs) is likely to minimize attrition in response to aggressive animals, but this rationale should be explained clearly to the owners present; violations of FIFO are generally perceived as being unfair and negatively impact the experience of those waiting in them.^41,42^ MDVC participant satisfaction should be prioritized wherever possible, as it impacts whether individuals will continue to participate in future MDVCs. Other behavioral interventions that can minimize queueing attrition is the use of messaging and incentives to flatten out the arrival rate. Field observations show arrival peaks, longer queue lengths, and greater attrition at midday (electronic supplementary materials, figure S3). Attrition during these peaks can be mitigated by communicating about shorter wait times early in the morning or incentivizing early arrivals by rewarding a limited quantity of “doorbuster” prizes (e.g., dog food or dewormer medication.

The expected vaccination coverage achieved by our optimization of fixed-location vaccination sites (57% and 48% for the low- and high-attrition scenarios, respectively) falls short of the 70-80% threshold recommended by WHO and PAHO.^37,43^ This gap can be met, in part, by combining fixed-location vaccination sites with mobile teams that deliver door-to-door vaccinations to areas with low penetration by the fixed-location campaign. This two-pronged approach has been leveraged successfully to achieve high vaccination coverage in other MDVCs^44,45^ as well as pandemic-era COVID-19 vaccination programs.^46,47^ A benefit of combining door-to-door vaccination with fixed-point vaccination is the ability to target high-risk or underserved areas, which not only increases total vaccine uptake but also promotes vaccine equity. We have previously found that the queue-naive algorithm increases the spatial evenness of vaccine coverage, a dimension of vaccine equity, even though it does not explicitly optimize for spatial equity.^26^ By placing more vaccination sites in more populous areas and limiting the placement of sites in less populous ones, the queue-conscious algorithm inadvertently decreases the spatial equity of fixed-point vaccinations compared to the queue-naive algorithm. In Arequipa, the less populous peri-urban areas also coincide with areas of greater socioeconomic disadvantage;^23,24^ thus, it is crucial for peri-urban areas to be prioritized by door-to-door campaigns following the deployment of fixed-point vaccination sites to ensure vaccine equity. This type of door-to-door outreach is particularly important for disadvantaged groups, who face the greatest barriers in accessing health services and are thus least able to travel to vaccination sites and wait for service.^48–50^ They might benefit the most from this combined approach.

There are several limitations of our study. The balking and reneging parameters *α* and *β* were not estimated from data but selected to model two hypothetical parameter regimes that fell within the upper and lower bounds of values that could feasibly capture real-world dynamics. While this lack of empirical estimation is a study limitation, our sensitivity analyses also indicated that the performance of our optimization algorithm was robust to misspecification of these parameters. In addition, the MDVC participation probability function that was used to optimize vaccination site locations included distance to the nearest site as a sole predictor and did not consider other household-level factors such as socioeconomic status (SES) or local environment factors such as urban/peri-urban status. Future studies can investigate how travel distance to MDVC sites affect MDVC participation among different household SES levels and across urban and peri-urban areas to derive a more nuanced MDVC participation function. Doing so can also be a means of promoting vaccine equity; for example, if future investigations revealed that marginalized groups are less able to travel long distances to participate in the MDVC, then the algorithm using this “updated” function would favor placing more sites near marginalized populations. Finally, our algorithm assumed that all MDVC sites were operated by a single vaccinator (*i.e.,* M/M/1). As a result, the algorithm tended to place multiple, adjacent single-vaccinator sites in highly populous areas. There are generally efficiency gains associated with multi-server (i.e., multi-vaccinator) queueing systems (where multiple vaccinators serve a single queue) compared to single-server systems with designated queues.^8^ However, pooling vaccinators (i.e., placing *k* vaccinators across fewer than *k* sites) may also lead to performance loss, as reducing the number of sites could result in longer queues, which may increase perceived waiting times and result in greater attrition;^51^ reducing the number of sites may also increase walking distances for some dog owners and thus decrease their probability of participation. A possible extension of our work would be to examine the tradeoff between gains from pooling vaccinators and losses due to slightly longer walking distances and potentially longer queue lengths.

In summary, our spatial optimization framework that incorporates expected losses from queueing offers insights for current vaccine-preventable disease programs and for future pandemic preparedness efforts. We developed a spatial optimization algorithm that maximizes total vaccine uptake by enhancing the spatial accessibility of vaccination sites while accounting for losses due to queueing attrition. We found that explicitly modeling queueing behavior, even with imprecise parameter estimates, led to gains in vaccination coverage and fewer losses to attrition than optimization that ignores the effects of queueing. Combined with door-to-door outreach and targeted media campaigns, rational placement of fixed-point vaccination sites is expected to bring vaccine uptake closer to threshold levels recommended for the control and eventual elimination of canine rabies. Considering the impact of excessive wait times on other vaccination campaigns, including the early rollout of the COVID-19 vaccine, our spatial optimization framework that explicitly considers queueing attrition can be broadly adopted to support other mass vaccination programs.

## Supporting information

Video animation of recursive interchange algorithm

## Data Availability

All code and non-sensitive data are publicly available on Github.

https://github.com/sherriexie/SpatialOptimizationQueueing

## DECLARATIONS

### Ethics statement

Ethical approval was obtained from the Institutional Review Board of Universidad Peruana Cayetano Heredia (approval number: 65369) and the University of Pennsylvania (approval number: 823736). All human subjects in this study were adults and informed consent was obtained from all subjects and/or their legal guardian(s). All methods were carried out in accordance with relevant guidelines and regulations.

### Data accessibility

All code and non-sensitive data are available on Github: https://github.com/sherriexie/SpatialOptimizationQueueing.

### Authors’ contributions

S.X., M.R., M.Z.L., and R.C.N. conceived of the project and designed the analyses. E.W.D. supervised the collection and curation of the data. S.X., M.R., and S.C. performed the analyses. S.X. drafted the manuscript and produced the figures. M.R., B.B., M.Z.L., and R.C.N. revised the manuscript and all authors read and approved the final manuscript.

## Acknowledgements

We gratefully acknowledge the contributions of and the work done by the Gerencia Regional de Salud de Arequipa, the Red de Salud Arequipa Caylloma, the Laboratorio Referencial Regional Arequipa, and the Microredes of the city of Arequipa. We acknowledge the work of the members of the Zoonotic Disease Research Laboratory, One Health Unit, and their contribution collecting part of the data used in this study.

