## Supplementary text and figures for "Enhancing Mass Vaccination Programs with Queueing Theory and Spatial Optimization"

**Electronic Supplementary Materials for**

### "Incorporating Queueing Theory into a Spatial Optimization Framework to Improve Mass Vaccination Programs"

### Supplementary Text A
### Steady-State Equations for M/M/1 FIFO Queueing System with Balking and Reneging

Our objective is to find closed-form equations to describe the behavior of a modified M/M/1 queueing model. In the following, we use the Markov chain $\{X_t,\ t \geq 0\}$ to denote the number of customers in the system at time $t$, i.e., customers either waiting in queue or being served. Balking is modeled via a modified arrival rate $\lambda_{s,n} = \lambda_s e^{-\alpha n/\mu}$ and reneging is modeled using modified service rate $\mu_n = \mu + (n-1)\beta$ whenever there are $n$ customers in the system. Note that $s\epsilon S$, where $S$ is the set of all selected vaccination sites, and the arrival rate $\lambda_s$ is allowed to vary across vaccination sites depending on the catchment size and the distribution of household travel distances for each site. However, the service rate $\mu$ (the inverse of the time it takes to administer a canine rabies vaccine) is expected to be constant across sites.

First, we note that $\{X_t,\ t \geq 0\}$ represents a birth-death process with rate $\lambda_{s,n}$ describing transitions from state $n$ to state $n+1$ and rate $\mu_n$ describing transitions from state $n$ to state $n-1$. A useful result given by equations (3.3) and (3.4) in *Fundamentals of Queueing Theory*[1] is that at equilibrium, the probability of finding $n$ customers (dogs) in a birth-and-death queueing process is equal to $p_n = \frac{\lambda_{n-1}\lambda_{n-2}\ldots\lambda_0}{\mu_n\mu_{n-1}\ldots\mu_1}p_0$ for $n \geq 1$ and $p_0 = \left(1 + \sum_{n=1}^{\infty} \frac{\lambda_{n-1}\lambda_{n-2}\ldots\lambda_0}{\mu_n\mu_{n-1}\ldots\mu_1}\right)^{-1}$.

Applying this result to the functional forms of $\lambda_{n,s}$ and $\mu_n$ specified above yields:

$$p_{s,n} = \frac{e^{-\frac{\alpha(n-1)}{\mu} - \frac{\alpha(n-2)}{\mu} - \ldots - \frac{\alpha}{\mu}}\lambda_s^n}{\prod_{i=1}^{n}(\mu + (i-1)\beta)}p_0$$

$$p_{s,n} = \frac{e^{-\frac{\alpha}{\mu}(1+2+\ldots+(n-1))}\lambda_s^n}{\beta^n \frac{\Gamma(n+\frac{\mu}{\beta})}{\Gamma(\frac{\mu}{\beta})}}p_0$$

$$p_{s,n} = \frac{e^{\frac{-\alpha n(n-1)}{2\mu}}\lambda_s^n \Gamma(\frac{\mu}{\beta})}{\beta^n \Gamma(n+\frac{\mu}{\beta})}p_0,$$

where $\Gamma(z)$ denotes the gamma function, i.e., $\Gamma(n) = (n-1)!$ for any integer $n > 0$ and $\Gamma(z) = \int t^{z-1}e^{-t}dt$ interpolates the factorial function to non-integer values. The probability $p_{s,0}$ is chosen so that $\sum_{n=0}^{\infty} p_{s,n} = 1$, and thus:

$$p_{s,0} = \left[1 + \sum_{n=1}^{\infty} \frac{e^{\frac{-\alpha n(n-1)}{2\mu}}\lambda_s^n \Gamma(\frac{\mu}{\beta})}{\beta^n \Gamma(n+\frac{\mu}{\beta})}\right]^{-1}$$

Note that the rate a which dogs join the queue (i.e., the effective arrival rate) is equal to the weighted sum of modified arrival rate $\lambda_{s,n}$ multiplied by the probability that the queue is in state $n$: $\sum_{n=0}^{\infty} p_{s,n}\lambda_{s,n}$, and the rate that dogs exit the queue before getting vaccinated (i.e., the reneging rate) is equal to the weighted sum of exit rate $(n-1)\beta$ multiplied by the probability that the queue is in state $n$: $\sum_{n=1}^{\infty} p_{s,n}(n-1)\beta$.

Therefore, the expected rate at which dogs are vaccinated at vaccination site $s$, after consideration of balking and reneging, is equal to:

$$v_s = \sum_{n=0}^{\infty} p_{s,n}\lambda_{s,n} - \sum_{n=1}^{\infty} p_{s,n}(n-1)\beta$$

and the expected number of dogs vaccinated across all vaccination sites $V$ is equal to:

$$V = \sum_{s \epsilon S} v_s t$$

Similarly, we can find the expected number of dogs lost to attrition through either balking or reneging from the above. Note that the balking rate at site $s$, which we will call $b_s$, is equal to the difference between the baseline arrival rate and the effective arrival rate for site $s$:

$$b_s = \lambda_s - \sum_{n=0}^{\infty} p_{s,n}\lambda_n$$

and hence the expected number of dogs lost to balking across all sites $B$ is equal to:

$$B = \sum_{s \epsilon S} b_s t$$

Finally, the reneging rate at site $s$, which we denote $r_s$, is equal to:

$$r_s = \sum_{n=1}^{\infty} p_{s,n}(n-1)\beta$$

and thus the expected number of dogs lost to reneging across all sites $R$ is equal to:

$$R = \sum_{s \epsilon S} r_s t$$

#### Supplementary Text B
#### The Hybrid Recursive Interchange-Genetic Algorithm for MDVC Site Optimization

**Overview of the hybrid algorithm**

The hybrid recursive interchange-genetic algorithm optimizes the placement of vaccination sites through a recursive swap-one-out strategy, also known as a recursive interchange algorithm (RI). Because the RI does not guarantee finding a global optimum, we combine it with a genetic algorithm (GA), which mimics the biological process of natural selection to improve the "fitness" of the solutions obtained. The hybrid algorithm cycles between the RI and GA until the stopping condition is reached: when two back-to-back rounds of optimization do not increase the expected number of dogs vaccinated beyond any of the previous rounds (Figure S2).

The first round of optimization with the hybrid algorithm proceeds with 1,000 independent iterations of the RI, where each iteration begins with a different randomly sampled starting set of vaccination sites. All subsequent rounds of the algorithm begin with the GA, which mates optimized "parent" solutions from the RI to obtain "offspring," which are then fed to the RI as subsequent starting sets. At the end of each round, the best solution and its associated score (expected number of dogs vaccinated) are retained to assess the stopping condition.

To facilitate the process of crossover in the GA, the study area was split into four zones (Figure S3a). The boundaries between the two zones in the north (Zones 1 and 2) and between Zone 2 and the zones in the south (Zones 3 and 4) follow the natural barriers formed by dry water channels that run across the area. The boundary between Zones 3 and 4 in the south were drawn to divide the region into zones of roughly equal populations. Detailed descriptions of the RI and GA are provided below.

**Recursive interchange algorithm**

The RI uses a recursive, swap-one-out strategy to find a locally optimal solution for a given starting set of vaccination sites (see the supplementary video). The objective function for the RI is the expected number of dogs vaccinated, which can be obtained one of two ways depending on whether queueing attrition is accounted for in the optimization:

- For **queue-conscious** optimization, the expected number of dogs vaccinated is equal to $V = \sum_{s \epsilon S} v_s t$, where $v_s$ is the queue-dependent vaccination rate at site *s* and *t* is the total time that each MDVC site operates (equations 5-6 in the main text).
- For **queue-naive** optimization, there is no queueing attrition so all dogs that arrive are assumed to get vaccinated. Thus the expected number of dogs vaccinated is simply equal to ***λt***, or the total dogs that arrive in time *t*.

The steps of the RI are as follows:

1. Specify the starting set to use in the RI, which will defer between the first and subsequent rounds of the hybrid algorithm
    - On round 1: Select a random set of 20 vaccination sites (out of the 70 candidate sites).
    - On round *n* (where n = 2, 3, 4, …): Start with a set of 20 vaccination sites that were obtained from the RI in the previous round of optimization.

2. For the starting set of sites, use the MDVC participation probability function to determine the expected number of arrivals at each site and divide this number by *t* (total operational time, which is assumed to equal 16 hours) to obtain the hourly arrival rate *λ* at each site.
3. Calculate the expected number of dogs vaccinated at each site (*Vt* for queue-conscious optimization and *λt* for queue-naive optimization) and sum across all sites to calculate the total number of dogs vaccinated.
4. Exchange one selected site with all non-selected candidate locations and keep the one that maximizes the total number of dogs vaccinated.
5. Repeat step 3 with remaining sites to obtain a locally optimized set of sites (each optimized set is treated as a *parental set* in the GA that is detailed below).
6. Perform steps 1-5 over 1,000 iterations to obtain 1,000 parental sets, which are subsequently used as input for the GA in the following round of optimization if the stopping condition has not been reached.

**Genetic algorithm**

A GA mimics the process of natural selection to find solutions to optimization problems with a large and complex search space. The GA employed in our hybrid algorithm includes the biologically inspired operations of crossover and mutation. The crossover step of our GA imitates the process of genetic recombination, which promotes diversity by combining sites from each parental set but also preserves spatial information in the parental sets by conserving the selection of sites in the same zone. Mutation is performed to introduce random changes to the offspring set to increase diversity in the offspring population. Mating pairs include pairings between two "optimized" parents that were output by the RI in the previous round and pairings between an optimized parent and a "random" parent that was generated using a spatially-structured random sampling approach (see subsection below for details). The latter pairing further increases the diversity of the offspring sets.

The steps of the GA are as follows:

1. Pair parental sets for mating. There will be 1,000 mating pairs:
    - Five hundred pairs will comprise two optimized parents.
    - Five hundred pairs will comprise one optimized parent and one random parent.
2. Perform <u>CROSSOVER</u> (Figure S2b): randomly sample two zones from Parent 1 and retain all sites from those zones; then obtain all sites of the remaining two zones from Parent 2.
3. Add/delete sites to ensure there are *k* = 20 sites total:
    - If total sites > 20 ... *remove* sites at random until 20 sites are left (all chosen sites have an equal probability of being removed).
    - If total sites < 20 ... *add* sites at random until 20 sites are left (all unchosen sites have an equal probability of being added).
4. Perform <u>MUTATION</u> (Figure S2c): each site has a 10% chance of mutating in which it is replaced by an unchosen site from the same zone.
5. The resulting 1,000 offspring sets serve as inputs (starting sets) for the RI in the current round of optimization.

**Generating random parental sets for mating**

The spatially-structured random sampling approach that was used to generate random parents consisted of the following two steps:

1. Sample four sites in each of the four zones (to obtain 16 sites total).
2. Sample four additional sites at random with equal probability (to obtain 20 sites).

#### Supplementary Text C
#### Characterizing the relationship between number of arrivals and MDVC site performance for different time-varying arrival densities

The closed-form equations for the MDVC queueing system that can be used to estimate the expected number of dogs vaccinated and losses to attrition are derived under the assumption of a constant arrival rate (Supplementary Text A and equations 1-6 in the main text). Thus, a different approach was required to estimate these performance metrics for time-varying arrival rates. The general approach employed was to estimate the expected number of dogs vaccinated and losses to attrition for a range of total arrival populations based on the median value obtained in repeated simulations.

Eight scenarios were considered: four time-varying arrival density functions (figure S4) at both low- and high-attrition values for α and β. Stochastic simulations were performed for each scenario over a range of total number of arriving dogs (8-2,000) as follows:

1. The time-varying arrival densities for a given function shown in figure S4 were multiplied by $N$ to determine the number of dogs arriving in each half-hour interval, and then that number was divided by 0.5 hr to determine the arrival rate $\lambda$ for each half-hour interval.
2. The arrival rates were used to generate arrival times for each half-hour interval by drawing interarrival times from an exponential distribution with rate $\lambda$.
3. The MDVC queueing system was simulated over four hours, beginning with zero dogs at time zero, to simulate a single day of the campaign, and the total number of dogs vaccinated and lost to attrition were recorded.
4. Steps 1-3 were repeated four times to simulate the outcome for the MDVC at a given site (to mimic the campaign, which takes place over four four-hour days for each fixed-point vaccination site).
5. Steps 1-4 (representing a single iteration of the simulation) were repeated 1,000 times.

The results of these simulations (medians and interquartile ranges) are plotted in figure S11. The median values for both the number of dogs vaccinated and lost to attrition were obtained from the simulations and were then fit using natural splines with seven degrees of freedom (figure S11).

These spline functions were used in place of the closed-form equations (equations 5-6) to estimate the number of dogs vaccinated and lost to attrition when the low- and high-attrition solutions were applied to the eight scenarios. For each configuration of vaccination sites (either the low- or high-attrition solution), the spline functions were used to transform the number of dogs that were expected to arrive at each site to the number vaccinated and the number lost to attrition.

#### Supplementary Figures

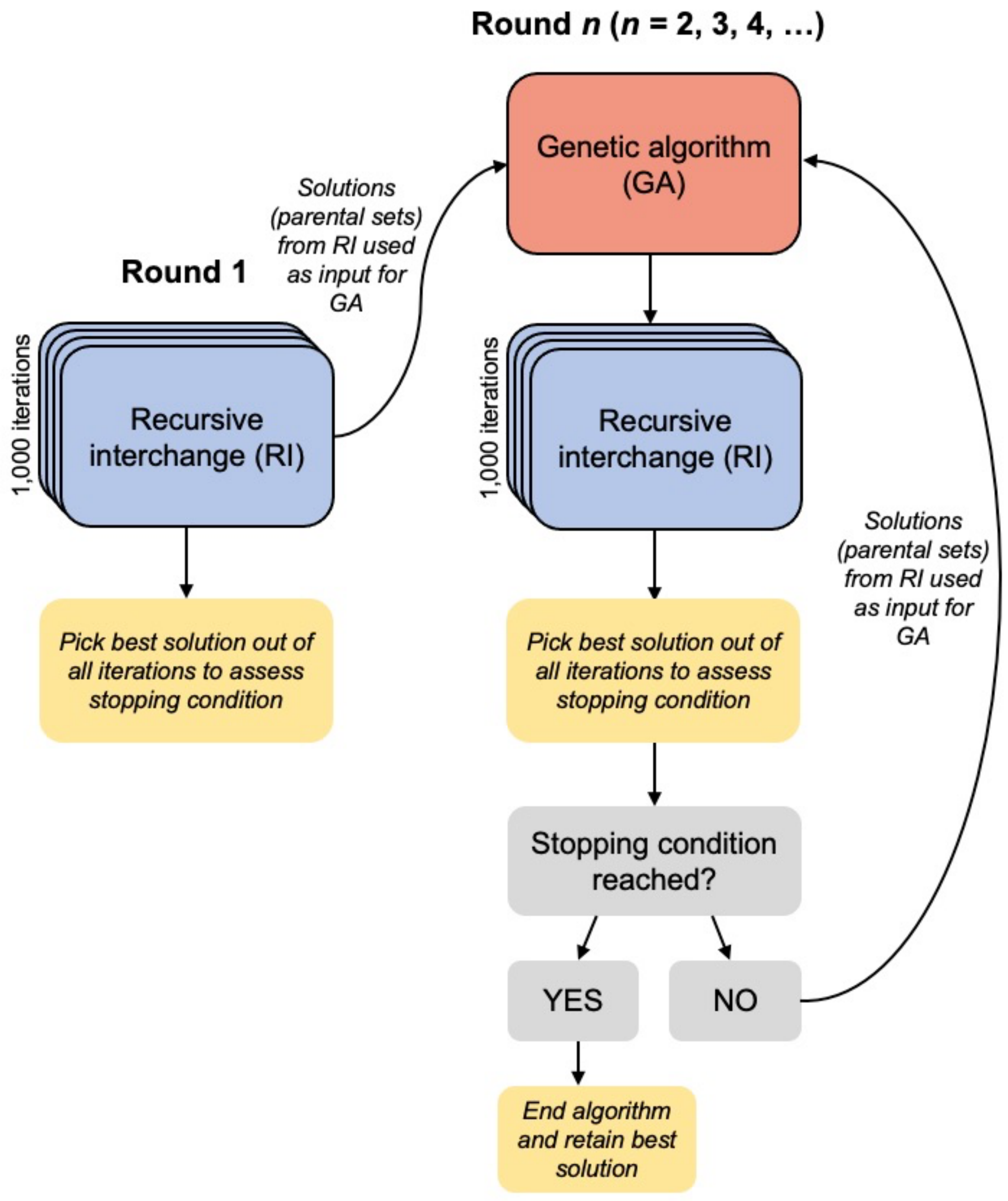

**Figure S1. Overview of the hybrid recursive interchange-genetic algorithm that was used to optimize the placement of MDVC sites.** The stopping condition for the algorithm is reached when the best solutions of the two latest rounds have a score (i.e., expected number of dogs vaccinated) that does not exceed the best score in all the previous rounds. More details about the algorithm can be found in the Methods section of the main text and Supplementary Text B.

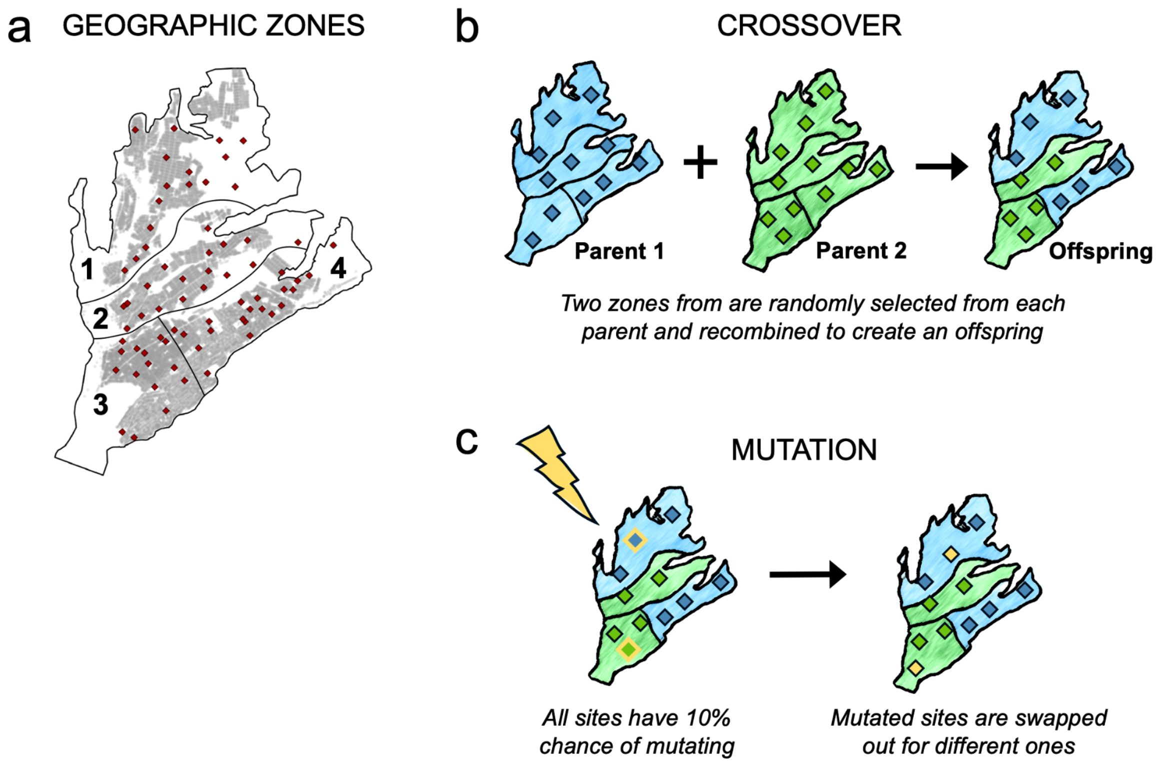

**Figure S2. Illustration of the genetic algorithm.** Panel a shows how the study area was divided into geographic zones to facilitate crossover. The seventy candidate sites are depicted as red diamonds, and the locations of houses are shaded gray. Panel b illustrates the process of crossover, in which two zones are randomly selected from each parent to generate an offspring. Panel c illustrates the process of mutation, in which all sites have a 10% change of mutating, and mutated sites are swapped with a non-selected site.

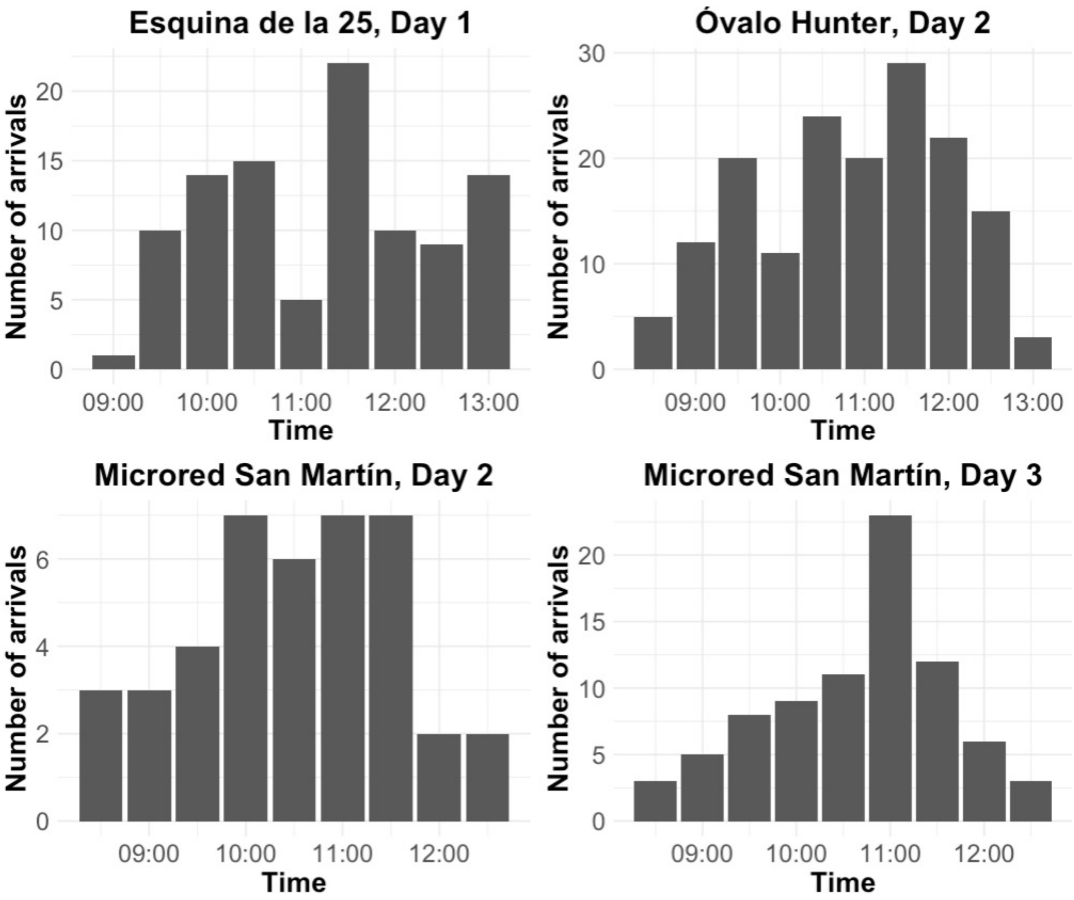

**Figure S3. Arrival times observed in the 2017 MDVC.** Arrival times observed at three different MDVC sites and on four separate days. The number of arrivals is binned in 30-minute intervals.

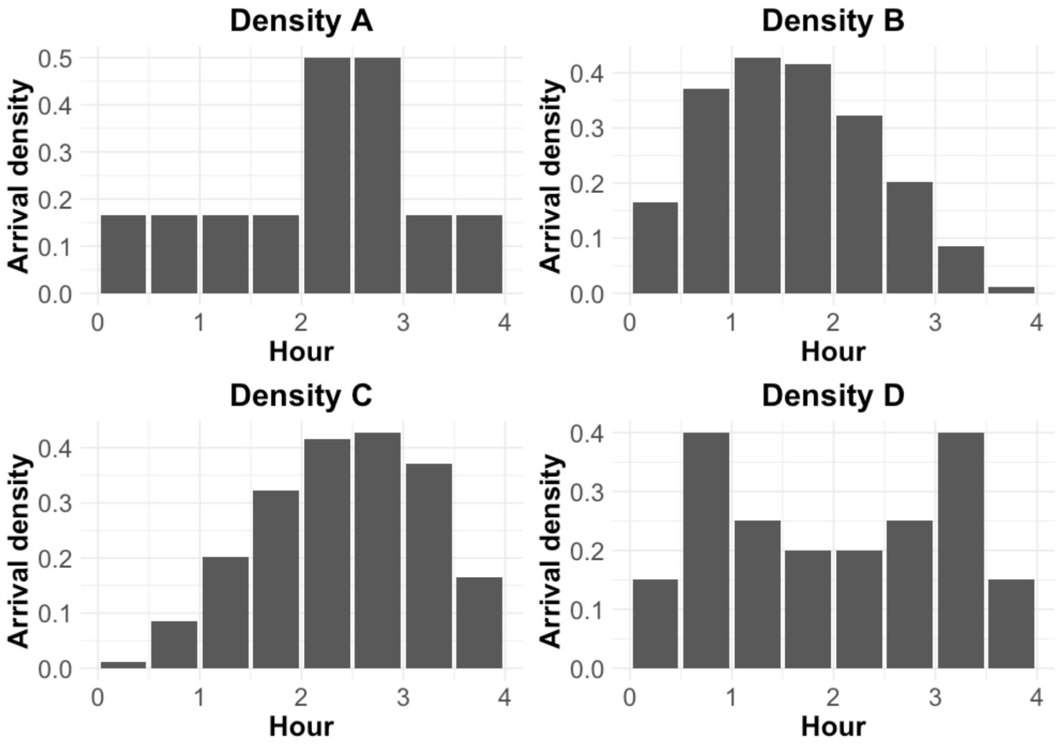

**Figure S4. Hypothetical time-varying arrival densities used to assess the relaxation of the constant arrival rate assumption.** The x-axis shows time progression over a 4-hour day at an MDVC site, and the y-axis gives the density of arrivals as the proportion of total arrivals for that day.

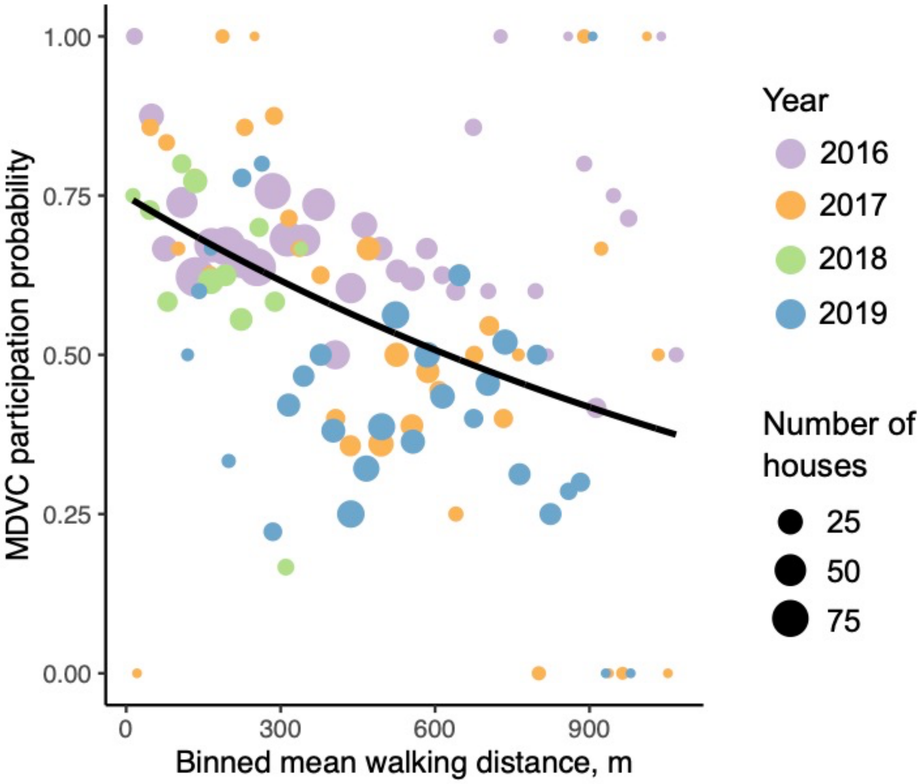

**Figure S5. The MDVC participation probability function.** Historical post-MDVC survey data are binned in 30-m distance increments to the closest MDVC site and visualized as dots that are colored by year and scaled by the number of houses per bin. The Poisson regression curve fit to survey data is shown by the solid, black line.

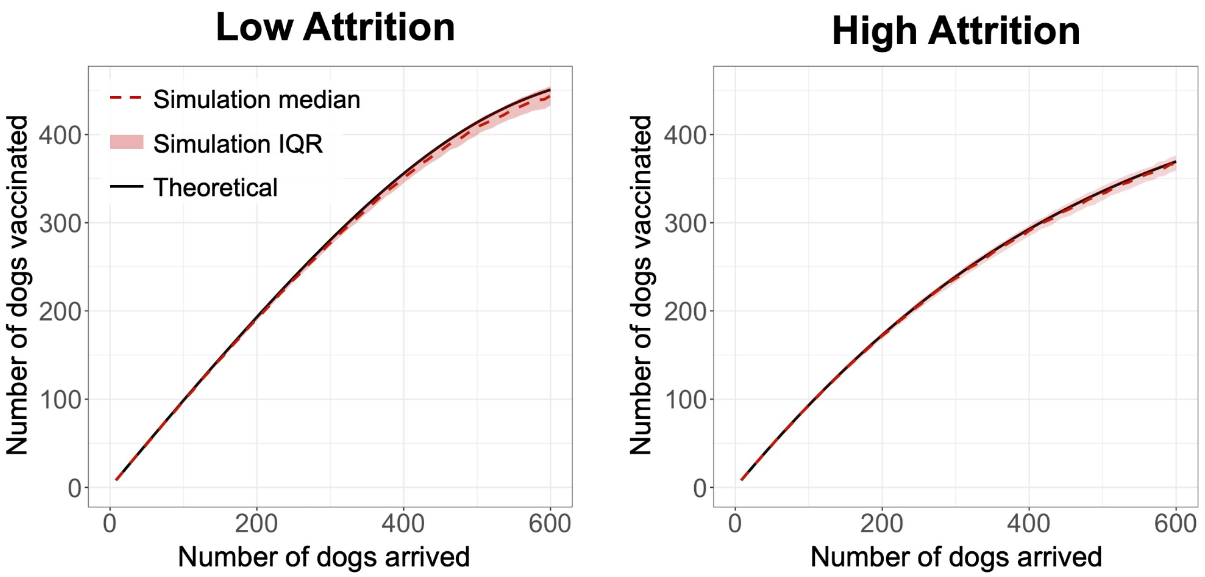

**Figure S6. Comparison of simulations with theoretical results.** The theoretical results obtained from equation 6 are shown by the solid black line, and simulation results are shown in red (median shown by the red, dotted line and the interquartile range shown by the red shaded area). Simulations were conducted for low- and high-attrition parameter regimes (low: $\alpha$ = 0.01 and $\beta$ = 0.02; high: $\alpha$ = 0.1 and $\beta$ = 0.1) and for a range of arrival rates (0.5-37.5 dogs/hour in increments of 0.5 dogs/hour or, equivalently, 8-600 total dogs arriving over 16 hours in increments of 4 dogs). The service rate $\mu$ = 30 $hr^{-1}$ for both the simulations and the input for equation 6. Each simulation consisted of four independent four-hour-long trials and compared to the output of equation 6 with $t$ = 16 hours. Simulations were repeated over 1,000 iterations for each set of parameter values.

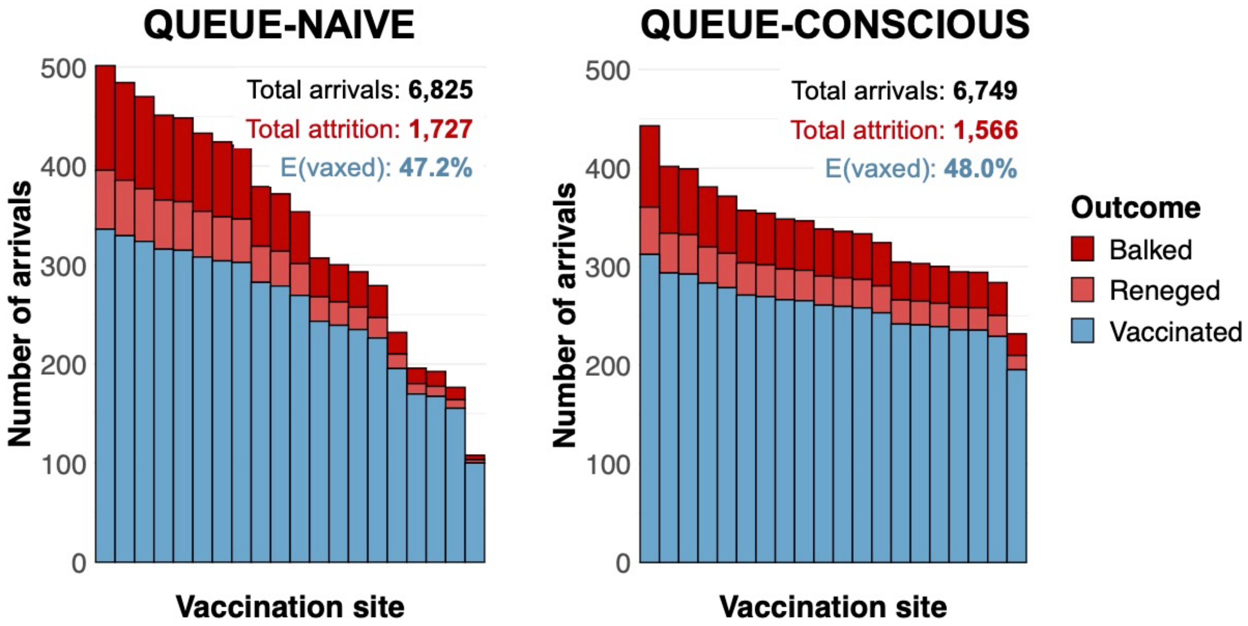

**Figure S7. Arrivals histograms for sites selected by queue-naive vs. queue-conscious optimization for the high-attrition system ($\alpha$ = 0.1, $\beta$ = 0.1).** The height of each stacked bar represents the expected number of dogs that arrive at a selected vaccination site. Bars are subdivided by color according to whether dogs ultimately get vaccinated (blue) or are lost to attrition, either through balking (dark red) or reneging (light red). The text above the bars give the total number of arrivals, total lost to attrition, and overall vaccination coverage achieved for each algorithm. While the queue-naive sites were obtained by the hybrid algorithm without considering attrition, the number of dogs vaccinated and the number of dogs lost to attrition for both queue-naive and queue-conscious sites were determined assuming high-attrition parameter values using equation 6 and the equations outlined in the Supplementary Text A.

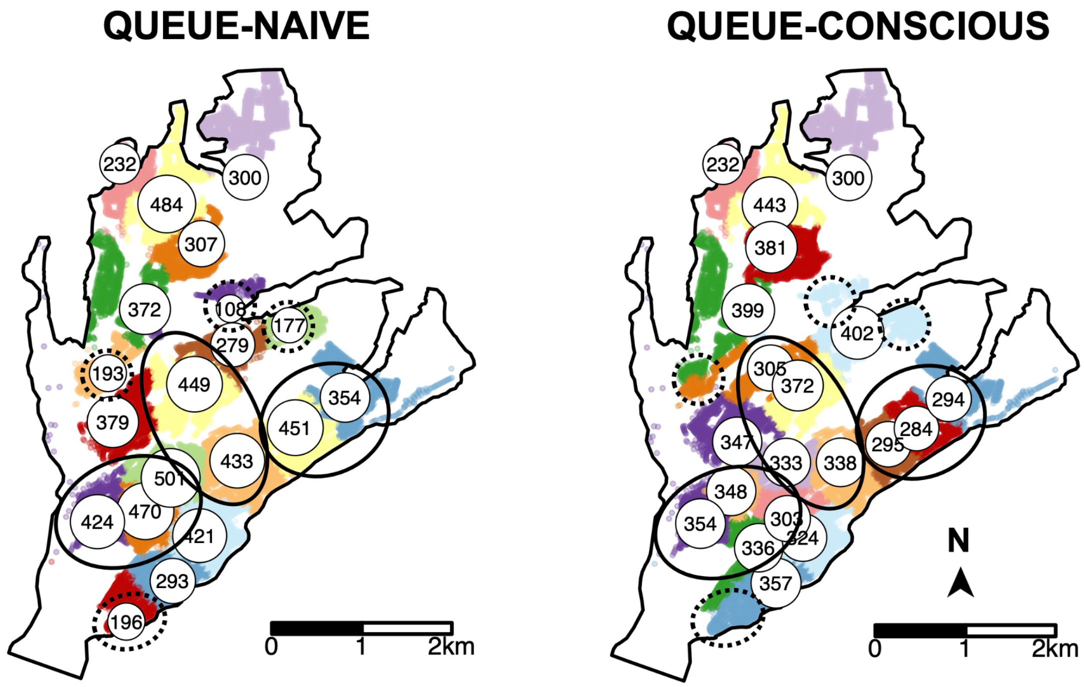

**Figure S8. Queue-naive vs. queue-conscious optimization for the high-attrition system ($\alpha$ = 0.1, $\beta$ = 0.1).** The locations of selected vaccination sites are indicated by white circles that are labeled and scaled according to the expected number of arriving dogs, which were calculated using equation 6. Houses in the study area are small dots colored according to their catchment, representing the area in which a MDVC site is the closest site for houses in terms of travel distance. Areas in which queue-conscious optimization concentrated more sites compared to queue-naive optimization are indicated by ellipses with solid lines, and areas in which queue-conscious optimization selected fewer sites are indicated by ellipses with dotted lines.

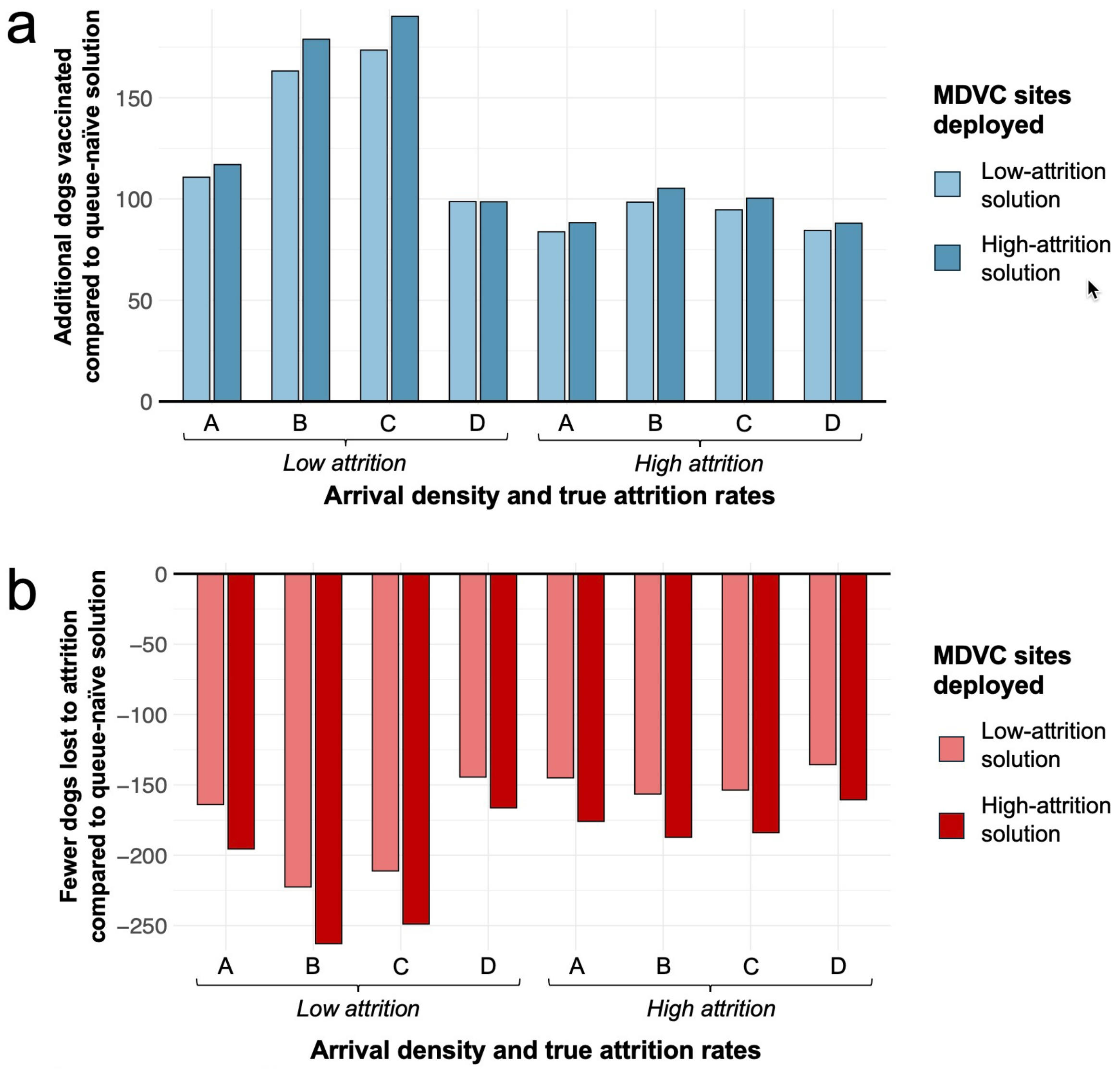

**Figure S9. Sensitivity of results to the constant arrival rate assumption: comparing queue-conscious vs. queue-naive optimization.** Panels illustrate how the low- and high-attrition solutions perform in the presence of different time-varying arrival rates compared to the queue-naive solution. Panel a shows the additional dogs vaccinated beyond the number achieved with the queue-naive solution, and panel b shows the reduction in attrition compared to the queue-naive solution.The time-varying arrival densities labeled A-D are shown in figure S4.

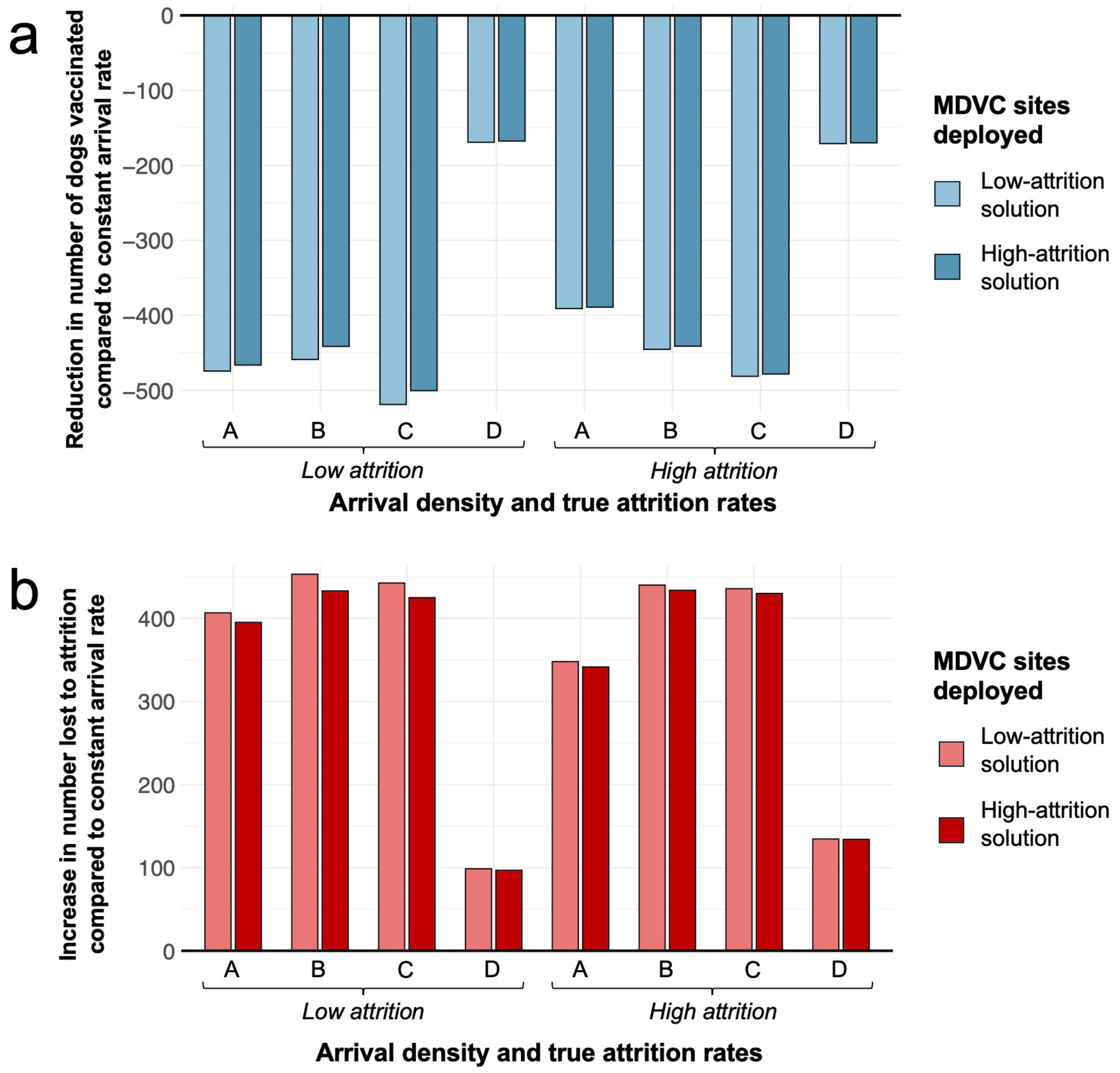

**Figure S10. Sensitivity of results to the constant arrival rate assumption: comparing estimates for constant vs. nonconstant arrival rates.** Panels illustrate how the low- and high-attrition solutions perform in the presence of different time-varying arrival rates compared to their performance when the constant arrival rate assumption holds. Panel a shows the reduction in the number of dogs vaccinated compared to when the arrival rate is constant, and panel b shows the increase in attrition compared to when the arrival rate is constant. The time-varying arrival densities labeled A-D are shown in figure S4.

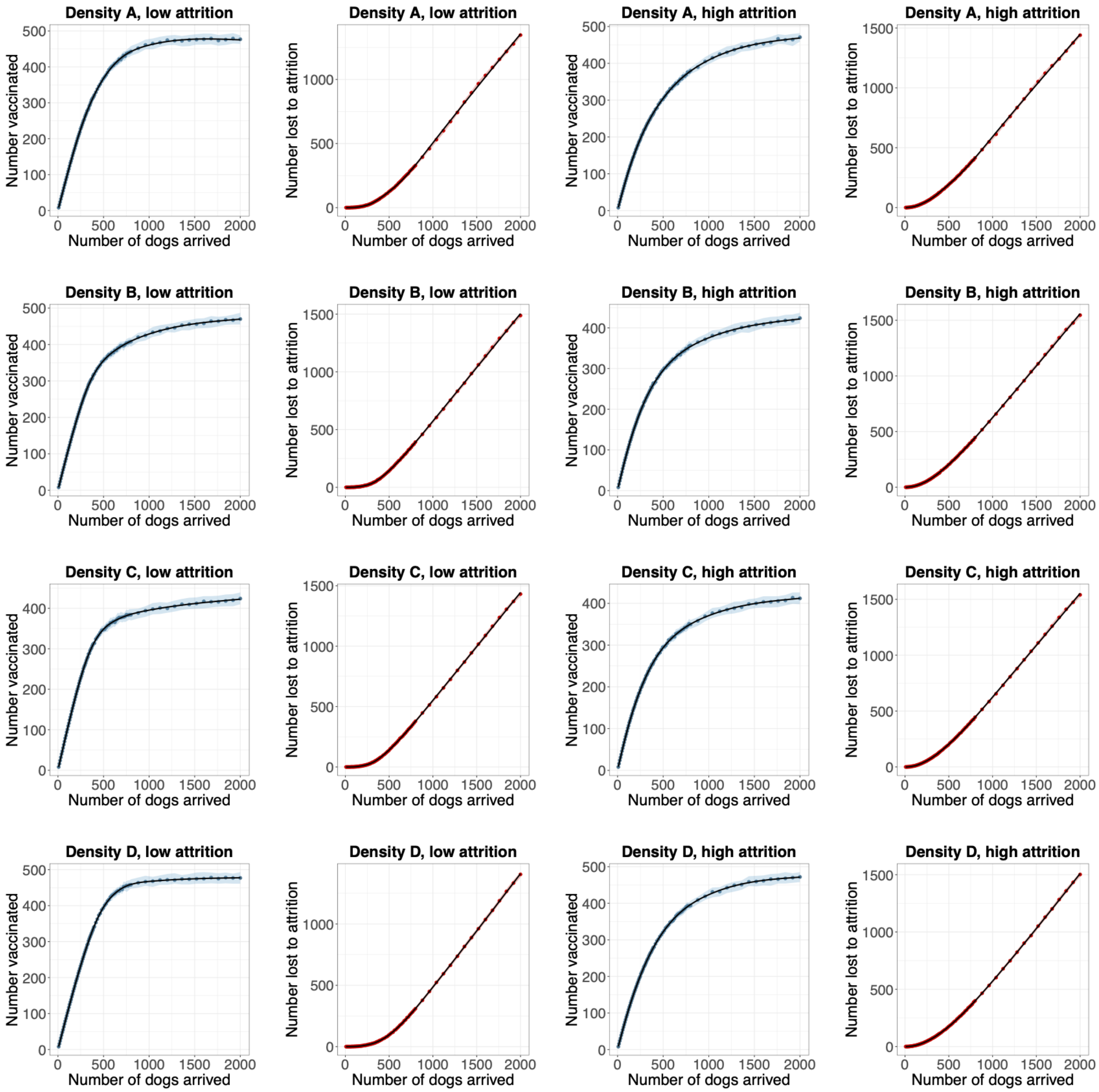

**Figure S11. Simulation results and regression splines for sensitivity analysis of constant arrival rate assumption.** Each plot shows either the number of dogs vaccinated or the number of dogs lost to attrition for a simulated campaign at an MDVC site. The simulations were run with one of the time-varying arrival rate densities shown in figure S4 and assuming either low- ($\alpha$ = 0.01 and $\beta$ = 0.02) or high-attrition ($\alpha$ = 0.1 and $\beta$ = 0.2) parameter values; the total number of dogs arrived was varied from 8-2,000 for each combination of arrival densities and attrition parameters. Each set of simulation conditions was repeated over 1,000 iterations. The median values obtained by each set of 1,000 iterations are shown by either the blue or red points, and interquartile ranges are shown by the shaded areas. The median values from the simulation form the basis for a continuous line approximation obtained by natural splines with seven degrees of freedom. These functions are shown as black lines and serve as input to the optimization model in the case of nonstationary arrival patterns.
